# Associations between Sympathetic Nerve Activity and Ischemia in Takotsubo Syndrome and Stable Ischemic Heart Disease

**DOI:** 10.1101/2024.01.10.24301128

**Authors:** Sanjana Borle, Xiao Liu, Anxhela Kote, Carine Rosenberg, Jewel Reaso, Peng-Sheng Chen, C. Noel Bairey Merz, Janet Wei

## Abstract

**Background:** Patients with stable ischemic heart disease (IHD) frequently experience ambulatory myocardial ischemia. Takotsubo Syndrome (TTS) often mimics IHD and is associated with heightened sympathetic activity. We hypothesized that skin sympathetic nerve activity (SKNA) differs between participants with prior TTS, stable IHD, and reference controls and that SKNA increases relative to electrocardiographic (ECG) evidence of ischemia.

**Methods:** Simultaneous recordings of SKNA and ECG (neuECG) measured ST height and average SKNA (aSKNA) in ambulatory participants with a history of TTS (n=6), stable IHD (n=7), and reference controls (n=19). Ambulatory ischemia was defined as ischemic ST segment depression ≥0.5 mm. SKNA burst frequency, duration, amplitude, and total area above the threshold were calculated during ischemic and non-ischemic episodes.

**Results:** Baseline aSKNA was similar between TTS, IHD, and reference controls (0.980± 0.061 µV vs 0.916±0.050 µV vs 1.098±0.291 µV respectively, p=0.06). Ischemic episodes (n=15) were identified in 2 TTS and 4 IHD participants, while reference controls had none. Among the TTS and IHD groups, ischemic ST depression was associated with increased heart rate and elevated aSKNA from baseline. An analysis of SKNA burst patterns at similar heart rates revealed that SKNA total burst area was significantly higher during ischemic episodes than non-ischemic episodes (0.301±0.380 µV.s and 0.165±0.205 µV.s, p=0.023) in both the TTS and IHD participants.

**Conclusions:** Compared with heart rate-matched non-ischemic episodes, ischemic episodes were associated with larger SKNA burst areas in the TTS and IHD groups. The neuECG may be a useful tool in studying the pathophysiology of sympathetic nervous system-related ischemia.

**Clinical Perspective:**

- Ambulatory monitoring with neuECG can assess sympathetic skin nerve activity (SKNA) and electrocardiography simultaneously.
- We found that ischemic episodes are present not only in patients with stable ischemic heart disease (SIHD) but also those with a history of Takotsubo Syndrome.
- SKNA burst activity was greater during ischemic episodes than nonischemic episodes in patients with SIHD and history of Takotsubo Syndrome.
- NeuECG may be a noninvasive tool in studying mechanisms of sympathetic nervous system-related ischemia.

## Introduction

Patients with stable ischemic heart disease (IHD) can exhibit signs of ambulatory myocardial ischemia, as noted by the depression of the ST segment on the electrocardiogram (ECG), which has been associated with a higher risk of cardiac mortality ^1^. Historical studies of ambulatory ischemia have demonstrated that while most episodes of ambulatory ischemia are associated with a preceding period of increased heart rate (HR), a minority of ischemic episodes are not associated with HR acceleration ^2,3^.

While the cardiac autonomic nervous system has long been implicated in the etiology of ambulatory ischemia, recent studies suggest that episodic coronary vasoconstriction may be responsible for the occurrence of ischemia in ambulatory patients. Microneurography recording of the muscle sympathetic nerve activity (SNA) has demonstrated increased activity during vasospastic angina, suggesting that SNA participates in the pathogenesis of vasospastic angina by enhancing coronary vascular tone ^4^. Indeed, ambulatory ischemia has been observed not only in patients with obstructive coronary artery disease (CAD) but also among those with signs and symptoms of ischemic heart disease (IHD) without obstructive CAD due to coronary vasospasm and/or microvascular dysfunction ^5^.

Takotsubo Syndrome (TTS), also known as stress cardiomyopathy, often mimics acute IHD and is associated with enhanced central and peripheral sympathetic stimulation ^6^. While mechanisms of acute TTS have been attributed to abnormalities of the SNA, sympathetic dysregulation is less well understood in patients who have recovered from TTS ^7^. Furthermore, patients with a history of TTS commonly experience chest pain, but it is not known whether they have evidence of ambulatory ischemia.

Currently, it is unknown whether elevated SNA in ambulatory patients with TTS or IHD is associated with ischemia. neuECG is a new method to simultaneously record ECG and skin sympathetic nerve activity (SKNA) in ambulatory patients ^8^ and has been validated by extensive canine and human subject studies ^9^. The purpose of the present study was to test the hypothesis that SKNA differs between stable TTS and IHD patients compared to reference controls, and that SKNA increases are associated with ECG evidence of ischemia.

## Methods

The study sample included patients with a history of TTS (n=6), stable IHD (n=7), and 19 reference controls. Reference controls were asymptomatic participants without known cardiovascular diseases and not taking prescription cardiac medications ^10^. All participants were provided a commercially available ECG monitor (Bittium FAROS 180, Oulu, Finland), which was placed vertically on the participant’s sternum for continuous ambulatory recording for 7 days. The vertical position was selected to capture a representative inferior ECG lead, given prior data indicating a high prevalence of ischemic ST depressions in the inferior leads during ambulatory monitoring among women with stable IHD ^5^. Seven participants underwent at least one repeat 7-day recording 46±21 days later. All medications including beta-blockers were continued during testing periods.

### Data Analyses

Ambulatory ECG and SKNA data were analyzed using the LabChart Pro 8 software. ECG data was filtered using a band pass of 0.5 to 150 Hz. A template previously established by the neuECG protocol was used to gather filtered SKNA, integrated SKNA (iSKNA), and average SKNA (aSKNA) ^10^. One-minute averages of ST segment height, SKNA, and heart rate (HR) were obtained from LabChart and analyzed as discrete data points. ST height was measured by LabChart at 110 ms after the QRS peak. An ischemic episode was defined as at least 30 seconds of horizontal or down-sloping ST segment depression ≥ 0.5 mm, when measured 80 ms after the J-point. A cardiologist manually verified all ischemic episodes. For ischemia burst analyses, each ischemic episode was also paired with a corresponding control episode defined as a non-ischemic episode that occurred within one hour of the ischemic episode and in which the average heart rate was similar to the ischemic episode ± 5 beats per minute (BPM). SKNA data during ischemic episodes were also analyzed separately and compared to corresponding control non-ischemic episodes using JMP Data Analysis Software. The software was used to identify the SKNA bursts using previously reported methods ^9^. A Microsoft Excel was used to calculate burst frequency (/min), duration (%), amplitude (µV), and total area above the threshold (µV) ^10^.

### Statistical Methods

Pearson regression analyses were performed to assess relationships between SKNA, HR, and ST segment height variables for each ambulatory recording. Student’s t-tests assuming unequal variance were used to compare the mean of means between two groups of participants, and single-factor ANOVA tests were used to compare between three or more groups. Shapiro–Wilk test was used to test for normality. Wilcoxon signed-rank test was used to compare the means if the data were not normally distributed. A P value ≤ 0.05 is considered significant.

### Study approval

The institutional review board of the Cedars-Sinai Medical Center approved a prospective observational study of participants undergoing ambulatory monitoring with neuECG. Written informed consent was obtained from all participants prior to participation.

### Data availability

The data supporting the findings of this study are available from the corresponding author upon reasonable request.

## Results

Table 1 shows the participant characteristics, comorbidities, and current medications. All TTS participants were female, and 3 (43%) of the IHD patients, who all had a primary diagnosis of obstructive CAD, were male. IHD participants with no obstructive CAD (n=4) were all female and had a suspected or confirmed clinical diagnosis of coronary microvascular dysfunction or vasospasm. Beta-blocker use was more frequent in the IHD group than TTS group. Five ischemic episodes were identified in 2 TTS participants, while 15 ischemic episodes were identified in 4 IHD participants. All ischemic episodes were asymptomatic. The reference control participants did not have ischemic episodes. The mean aSKNA was 0.980 ± 0.061 µV for all TTS recordings, 0.916 ± 0.050 µV for IHD recordings, and 1.098 ± 0.291 µV for reference control recordings (p=0.06).

**Table 1.** Participant characteristics, comorbidities, and current medications. IHD, ischemic heart disease; TTS, Takotsubo syndrome; *, p<0.0001 between groups by ANOVA

| Mean (range) or N (%) | **TTS**<br>N=6 | **IHD**<br>N=7 | **Reference Control**<br>N=19 |
| --- | --- | --- | --- |
| **Age, years\*** | 65 (62-78) | 61 (26-76) | 30 (20-48) |
| **Sex, female** | 6 (100%) | 4 (57%) | 19 (100%) |
| **Hypertension** | 4 (67%) | 4 (57%) | 0 (0%) |
| **Hyperlipidemia** | 3 (50%) | 3 (43%) | 0 (0%) |
| **Diabetes** | 0 (0%) | 2 (29%) | 0 (0%) |
| **Active Smoking** | 0 (0%) | 0 (0%) | 0 (0%) |
| **Coronary Artery Disease** | 0 (0%) | 3 (43%) | 0 (0%) |
| **Beta-Blocker** | 1 (17%) | 5 (71%) | 0 (0%) |
| **Calcium Channel Blocker** | 3 (50%) | 2 (29%) | 0 (0%) |
| **Nitroglycerin** | 4 (67%) | 4 (57%) | 0 (0%) |
| **Statin** | 2 (33%) | 3 (43%) | 0 (0%) |
| **Ivabradine** | 0 (0%) | 0 (0%) | 0 (0%) |
| **Benzodiazepine** | 0 (0%) | 0 (0%) | 0 (0%) |
| **Depression Medication** | 0 (0%) | 1 (14%) | 1 (5%) |

### A negative correlation between aSKNA and ST height

We found a significant negative correlation between aSKNA and ST height in all groups. **Figure 1** shows an example from a TTS participant. Figure 1A shows the changes of aSKNA, HR and ST height over time. Figure 1B shows a negative correlation between ST height and aSKNA and a positive correlation between HR and SKNA. The original recordings at points C-E in Panel B are shown in Panels C-E, respectively. These graphs show that low aSKNA was associated with normal ST segment (Panel C) while heightened aSKNA was associated elevated HR and ischemic ST depression (Panels D-E). **Figure 2** shows a similar analysis in an IHD participant. **Figure 2A** shows the aSKNA, ST height and HR over time. **Figure 2B** shows a strong negative correlation between aSKNA and ST. As in **Figure 1**, low aSKNA was associated with normal ST segment (**Figure 2C**). The ischemic ST segment depressions were associated with increased HR and elevated aSKNA (**Figures 2D and 2E**). **Figure 3** shows an example of upsloping ST depression in a reference control participant. There was a modest negative correlation between aSKNA and the ST height (**Figure 3B**). As expected, the recordings from a reference control participant did not have any ST depression indicating myocardial ischemia either at either low (**Figure 3C**) or high SKNA (**Figure 3D**).

**Figure 1.**
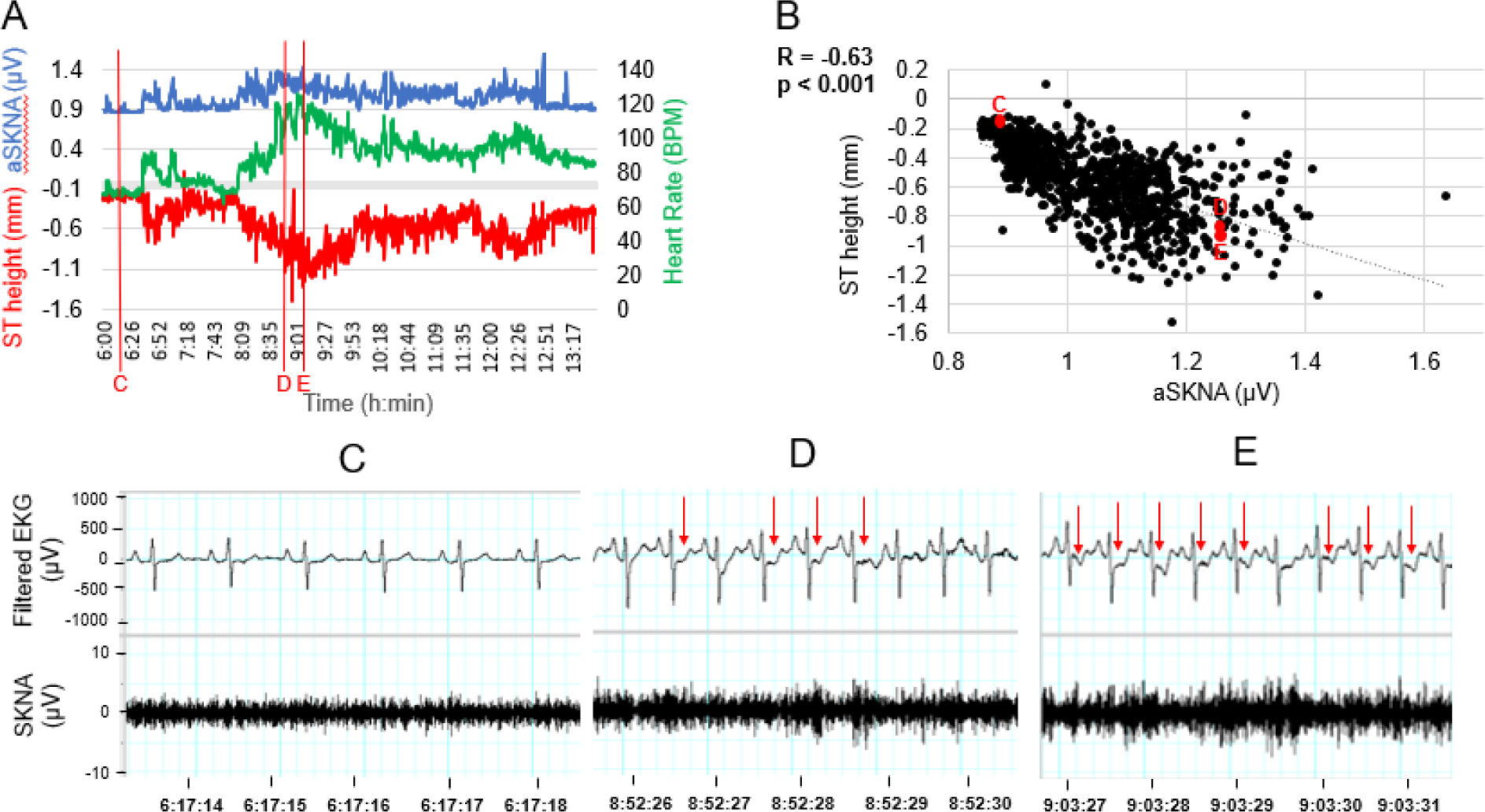
Temporal changes of aSKNA, ST height, and HR in an ambulatory 78-year-old female participant with a history of Takotsubo Syndrome. A 7-hour segment of her recording was analyzed min-by-min for aSKNA, Heart Rate, and ST height values. The data were plotted over time, showing that elevated aSKNA is associated with increased HR and depressed ST (A). B shows a regression between aSKNA and ST height during this period. The original recordings at points C-E in B are shown in Panels C-E, respectively. Arrows in D and E point to ischemic ST segment depression, which occurred during heightened SKNA.

**Figure 2.**
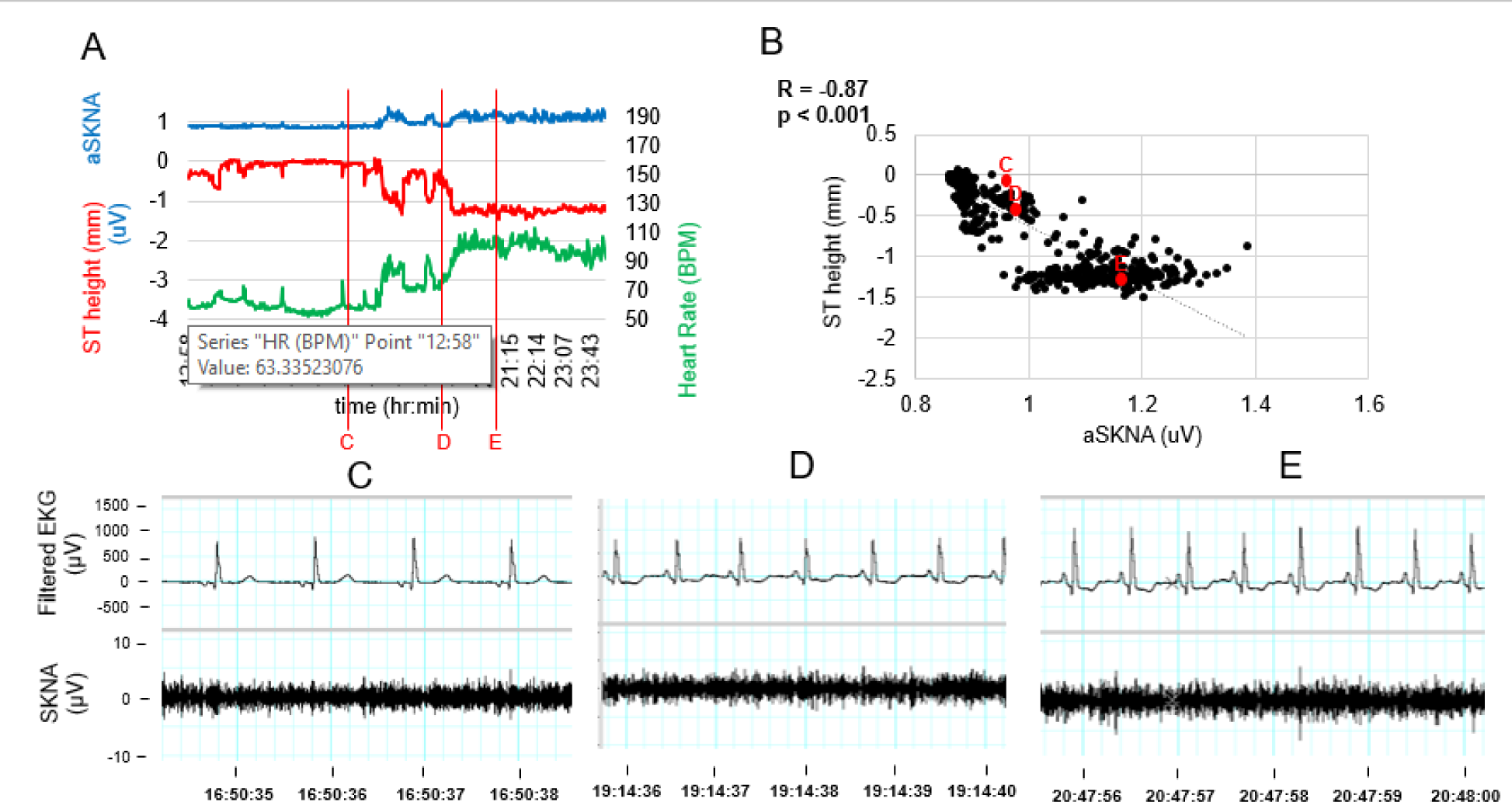
Temporal changes of aSKNA, ST height, and HR in a 63-year-old woman with ischemic heart disease but no obstructive CAD. A shows the changes over 11 hours. B shows a negative correlation between aSKNA and ST height. The original recordings at points C-E are shown in Panels C-E, respectively. C shows an absence of ST depression when aSKNA was low. Arrows in D and E point to ischemic ST segment depression, which occurred during heightened aSKNA.

**Figure 3.**
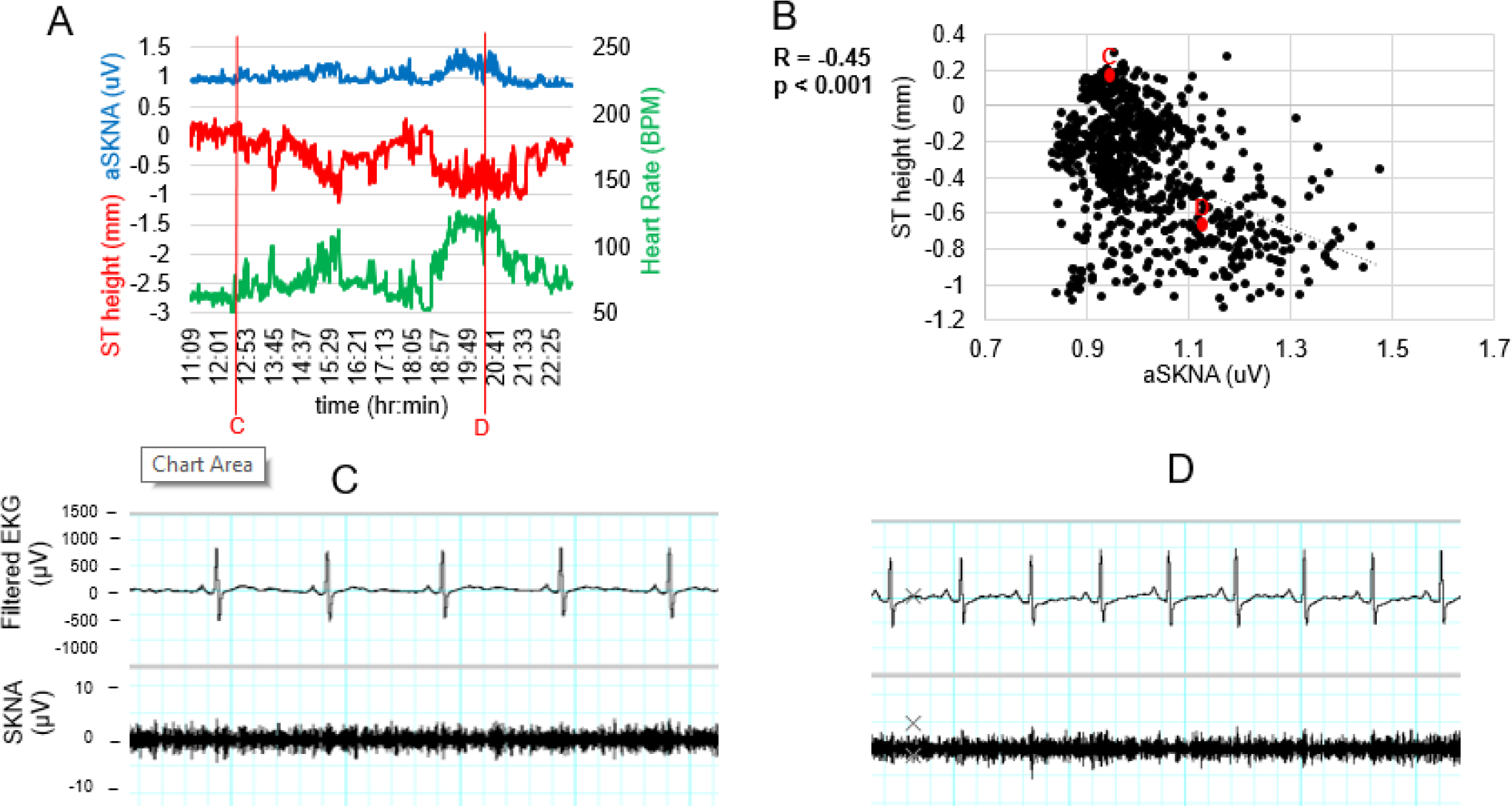
Temporal changes of aSKNA, ST height, and HR in a 33-year-old female control participant. Numerical values for aSKNA, HR, and ST height values were averaged min-by-min and plotted over the course of an 11-hour recording segment (A). B shows regression between aSKNA and ST height. The original recordings at points C and D are shown in Panels C and D, respectively. No ischemic ST depression was found in this or any other control participants.

### Ischemic and non-ischemic episodes of similar heart rate

The difference in SKNA between non-ischemic and ischemic episodes at similar heart rates, as suggested by the previous results, was further investigated by conducting a thorough SKNA burst analysis (**Figure 4**). The ischemic ST segment depression (**Figure 4A**) was associated with more frequent SKNA bursts and larger total burst area than the non-ischemic episode with similar heart rate (**Figure 4B**). We compared the 15 identified ischemic episodes in 6 participants (2 TTS and 4 IHD) to the 15 non-ischemic episodes with similar HR but no ischemic ST depression. The mean SKNA burst area of ischemic and non-ischemic episodes were 0.301±0.38 µV.s and 0.165±0.205 µV.s, respectively (N=6, p=0.023 by Wilcoxon signed-rank test).

**Figure 4.**
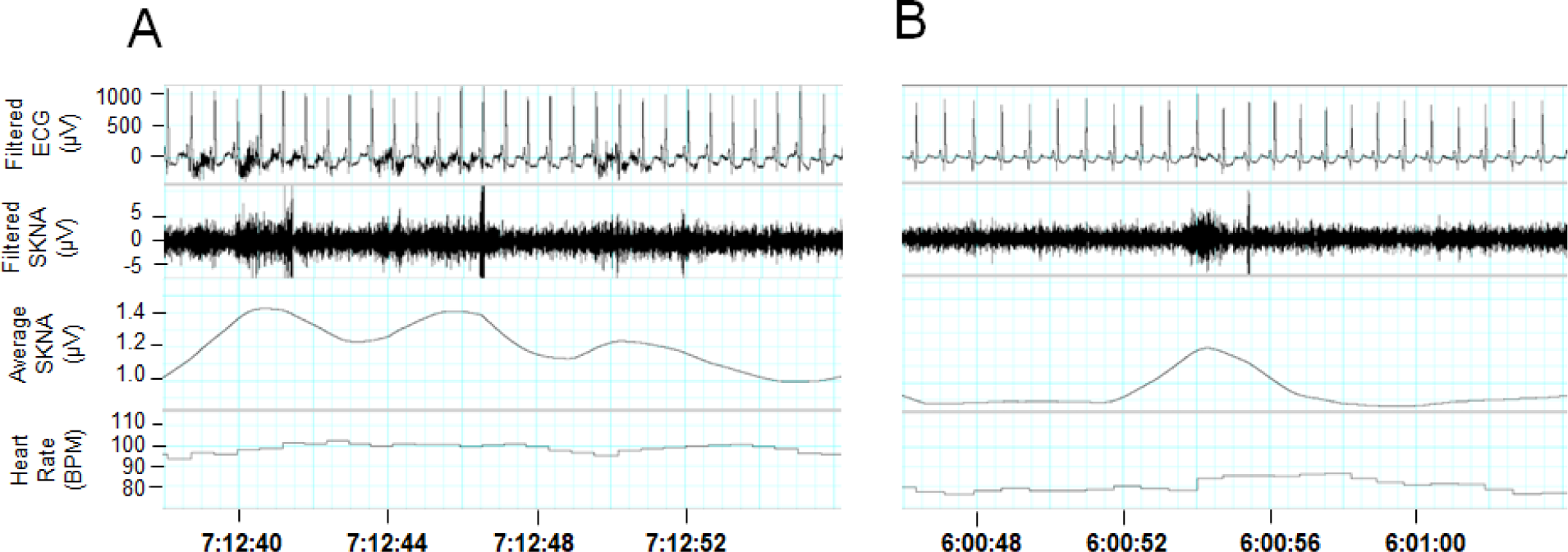
Differences of aSKNA burst area between ischemic and non-ischemic episodes in the same patient. This example was from a 63-year-old woman with IHD. We first identified an episode with ischemic ST segment depression (A). We then reviewed the same recording to find an episode with a similar heart rate but no ischemic ST segment depression (B). The solid red line on the aSKNA tracings indicates the burst threshold for that episode. The area above the threshold is the burst area. Note that there is a higher burst threshold and larger burst area in ischemic than in non-ischemic episodes.

## Discussion

We found similar aSKNA levels in TTS, IHD and reference controls, contrary to our hypothesis. Compared with heart rate-matched non-ischemic episodes, ischemic episodes were associated with larger SKNA burst areas in the IHD and TTS groups.

### The relationship between SKNA and ST depression

We found a negative correlation between aSKNA and ST height in all groups of patients. One potential mechanism for this observation is that sympathetic activity increased HR, which in turn caused repolarization changes that manifest as ST depression. However, a previous study showed that atrial pacing alone with rates of 120-180 bpm does not cause ST depression or coronary sinus oxygen desaturation in patients without coronary artery diseases ^11^. In comparison, adrenalin infusion can cause dose-dependent ST depression in all patients, but a great magnitude of depression was seen in patients with ST depression during exercise tests than in controls ^12^. In another study, the ST depression in patients with abnormal ST-T waves at baseline but without obstructive CAD is worsened by the combined use of atrial pacing and adrenalin ^13^. These findings suggest that sympathetic tone is the primary determinant of ST depression but that elevated HR could be a contributing factor in patients with pre-existing heart diseases.

### ST depression in asymptomatic reference controls

We found a negative correlation between ST height and aSKNA in reference control participants. However, none of the ST depressions were ischemic depressions. ST depression occurs when the junction (J) point is displaced below baseline ^14^. The J point correspond to the junction between QRS and ST segment. Its magnitude and timing are determined by the transmural dispersion of the transient outward current (I_to_). A prominent Ito-mediated action potential notch in ventricular epicardium but not endocardium produces a transmural voltage gradient during early ventricular repolarization that registers as a J wave or J-point elevation on the ECG ^15^. Transient outward channels are subject to α- and β-adrenergic regulation, mainly decreasing I_to1_ ^16^. It is possible that elevated SKNA suppressed the I_to_, which downwardly displaced the J point. This could be the mechanism of upsloping ST depression. All our control participants are women, who normally have lower magnitudes of I_to_ than men and thus may be more prone to J point depression during sympathetic stimulation. That may be another reason that prominent upsloping ST depression was observed during SKNA bursts.

### ST depression during ischemia

We found a negative correlation between ST height and aSKNA also in patients with TTS and IHD. Similar to the asymptomatic reference controls, majority of the ST depression in these diseased patients occurred in the setting of upsloping ST depression. However, with manual analyses, we were able to find 15 episodes in 6 patients of horizontal or down-sloping ST depression that cannot be explained simply by J point deviation. Those episodes were associated with significantly higher aSKNA levels than HR-matched control non-ischemic segments of the same patients. These findings suggest that those horizontal or down-sloping ST depression cannot be explained by the HR elevations. Rather, sympathetic stimulation increased myocardial ischemia, which caused ST depression.

### Therapeutic implications

In addition to obstructive CAD, myocardial ischemia can be caused by coronary vasospasm or coronary microvascular dysfunction, namely in patients with ischemia and no obstructive CAD^17^. Coronary vasospasm and coronary microvascular dysfunction have also been implicated in the pathophysiology of TTS ^7^. Since ischemic and non-ischemic episodes were matched for similar time of day, it is unlikely that circadian variation in ischemic activity would explain these findings ^18^. Both TTS and IHD groups demonstrated an overall increase in total burst area during ischemic episodes, even while most of them were on a beta-blocker as seen in **Table 1**. This suggests the potential relatively significant effects of autoregulation on ischemic episodes in these patient populations. Previous literature has suggested a future potential for using vagal stimulation to relieve severity of myocardial ischemia, and this study supports the hypothesis that reducing sympathetic activation may be beneficial for reducing myocardial ischemia ^19^.

## Limitations

Though this study offers promising preliminary results, it has some limitations. Because the sample size for both TTS and IHD participants was small, further observational analysis in these populations is warranted to further establish a clear relationship between sympathetic nerve activity and ischemia. It is also important to note that in this study, participants did not withhold any medications, and these might have affected ambulatory ECG and SKNA readings. The ECG and SKNA readings in this study also had varying levels of artifact from one participant to another, thus each recording did not have a consistent amount of clean, analyzable data. Furthermore, ECG recordings were available only in one lead, which reduces the sensitivity of ischemia detection.

## Conclusions

Compared with heart rate-matched non-ischemic episodes, ischemic episodes were associated with larger SKNA burst areas in the TTS and IHD groups. The neuECG may also be a useful tool in studying the pathophysiology of sympathetic nervous system-related ischemia.

## Author contributions

PC, NBM, and JW contributed to the design and supervision of the research. AK, CR, and JR acquired the data. SB, XL, PC, JW analyzed the data. SB, PC, NBM, and JW wrote the paper with input from all authors.

## Data Availability

Data available upon request.

## Acknowledgements

This study was supported in part by NIH Grants R01HL139829, OT2 OD028190, an AHA award 23IPA1052289, R01HL146158, U54AG065141, the Burns & Allen Chair in Cardiology Research, the Barbra Streisand Women’s Cardiovascular Research and Education Program, the Linda Joy Pollin Women’s Heart Health Program, the Erika Glazer Women’s Heart Health Project, and the Adelson Family Foundation, Cedars-Sinai Medical Center, Los Angeles, California.

